# Beta-triggered adaptive deep brain stimulation during reaching movement in Parkinson’s disease

**DOI:** 10.1101/2022.12.20.22283430

**Authors:** Shenghong He, Fahd Baig, Anca Merla, Flavie Torrecillos, Andrea Perera, Christoph Wiest, Jean Debarros, Moaad Benjaber, Michael G Hart, Francesca Morgante, Harutomo Hasegawa, Michael Samuel, Mark Edwards, Timothy Denison, Alek Pogosyan, Keyoumars Ashkan, Erlick Pereira, Huiling Tan

## Abstract

Subthalamic nucleus (STN) beta-triggered adaptive deep brain stimulation (ADBS) has been shown to provide clinical improvement comparable to conventional continuous DBS (CDBS) in people with Parkinson’s disease (PD) with less energy delivered to the brain and less stimulation induced side-effects. However, several questions remain unanswered. First, there is a normal physiological reduction of STN beta band power just prior to and during voluntary movement. ADBS systems will therefore reduce or cease stimulation during movement and could therefore compromise motor performance compared to CDBS. Second, beta power was smoothed and estimated over time periods of 400ms or longer in most previous ADBS studies. A shorter smoothing period could have the advantage of being more sensitive to changes in beta power which could enhance motor performance. In this study, we addressed these two questions by evaluating the effectiveness of STN beta-triggered ADBS using a standard 400ms and a shorter 200ms smoothing window during reaching movements. Results from 13 people with PD showed that STN beta-triggered ADBS is effective in improving motor performance during reaching movements as it better preserves gamma oscillation than CDBS in people with PD, and that shortening the smoothing window does not result in any additional behavioural benefit. ADBS significantly improved tremor compared with no DBS but was not as effective as CDBS. When developing ADBS systems for PD, it might not be necessary to track very fast beta dynamics; combining beta, gamma, and motor decoding might be more beneficial with additional biomarkers needed for optimal treatment of tremor.

## Introduction

Deep brain stimulation (DBS) targeting the subthalamic nucleus (STN) has been demonstrated to be a successful treatment for patients with advanced Parkinson’s disease (PD) (*1*). However, continuous DBS (CDBS) can reduce in efficacy over time and may be accompanied by stimulation related side-effects such as dyskinesia, postural instability, impairment of cognition and reduced speech fluency (*2, 3*).

Enhanced synchronisation of beta activity in the STN has been consistently observed in people with PD, and is positively correlated with bradykinesia and rigidity. Conversely, improvement in bradykinesia and rigidity with medication or DBS is positively correlated with suppression of beta power (*4-9*). More recently, multiple studies have emphasized the importance of the temporal dynamics of STN beta oscillations, where the occurrence of longer beta bursts are positively correlated with motor impairment (*10-13*). Taken together, these findings suggest that STN beta activity is a biomarker for parkinsonian motor symptoms, and has motivated the development of beta-triggered adaptive DBS (ADBS) algorithms. The results of several pilot trials of ADBS with temporarily externalized DBS electrodes (*8, 14-18*), or chronically implanted DBS devices (*19-21*) suggest that beta-triggered ADBS is at least as effective as conventional CDBS in reducing motor symptoms at rest as evaluated by Movement Disorders Society Unified Parkinson’s Disease Rating Scale (MDS-UPDRS) part III.

However, several questions remain unanswered. First, there is a physiological reduction of STN beta activity during voluntary movements, which is seen also in people with PD (*22-24*). In the setting of beta-triggered ADBS, this will lead to reduction or cessation of stimulation during movement. This could compromise motor performance compared with CDBS if further beta suppression during movement is helpful for maximum therapeutic benefits when patients attempt movements, which is arguably when they need it most (*25*). Second, most existing studies of beta-triggered ADBS have estimated beta amplitude in real-time using an average moving window of 400-millisecond duration or longer, aimed at capturing beta bursts of longer durations (*10, 14-16*). It is possible that there would be benefits from an ADBS algorithm capable of tracking faster beta dynamics using a shorter smoothing time window (e.g., 200ms).

To answer these questions, we developed an experimental protocol combining a cued reaching task and a brain computer interface allowing real-time estimation of STN beta and adjustment of stimulation parameters (**Fig. 1**). We evaluated the motor performance of 13 people with PD in four different stimulation conditions: no DBS, CDBS, ADBS-400 (ADBS with beta amplitude smoothed over 400ms), and ADBS-200 (ADBS with beta amplitude smoothed over 200ms).

**Figure 1.**
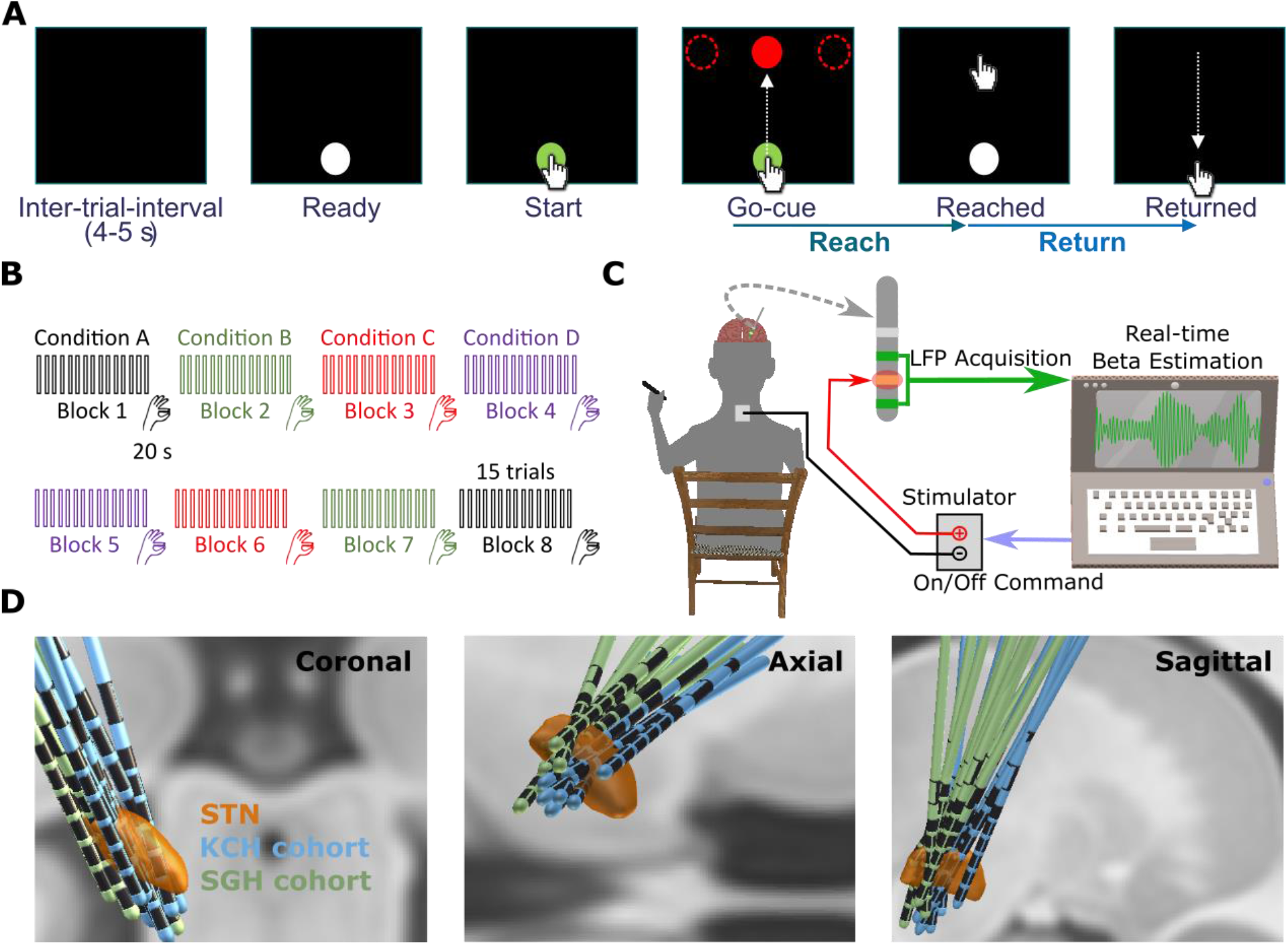
Experimental protocol. **(A)** Timeline of one individual trial of the reaching task performed on a tactile monitor with a pen. In each trial, patient is instructed to point at the start button to initiate the trial, reach to the red target when the Go-cue is shown, and back to the start button when the target disappears, as quickly as possible. **(B)** Timeline for the whole experimental session which consists of eight counterbalanced blocks in four different stimulation conditions, with two blocks in each condition. Each block contains 15 trials of reach-return movements followed by 20 second of finger-tapping movements. **(C)** Schematic of the adaptive DBS system which consists of bipolar measurement of subthalamic nucleus (STN) local field potentials (LFPs), real time estimation of beta amplitude, and monopolar stimulation delivered to one of the middle contacts while the patient is comfortably seated on a chair and performs the tasks. **(D)** Three-dimensional reconstruction in coronal (left), axial (middle), and sagittal (right) views of all analysed DBS leads localized in standard MNI-152_2009b space (*53*) using Lead-DBS (*26*). Electrodes in the left hemisphere were mirrored to the right hemisphere. Please see Supplementary Video 1 for a 360-degree view of the reconstruction results.

## Results

### 1. ADBS-200 led to more frequent stimulation switches and shorter beta burst durations, but no difference in motor performance compared with ADBS-400

To evaluate how different smoothing windows for calculating the beta amplitude may affect the performance of the ADBS, we first identified each stimulation switching event. As shown in **Fig. 2A**, as expected, the stimulation was overall switched on and off more frequently during ADBS-200 compared with ADBS-400 (*t*_15_ = 16.5321, *p* = 4.8823e-11, paired *t*-test, **Fig. 2A**), with no significant difference in the average percentage of time when the stimulation was switched on between the two ADBS conditions (*t*_15_ = -2.1327, *p* = 0.050, paired *t*-test, **Fig. 2B**).

**Figure 2.**
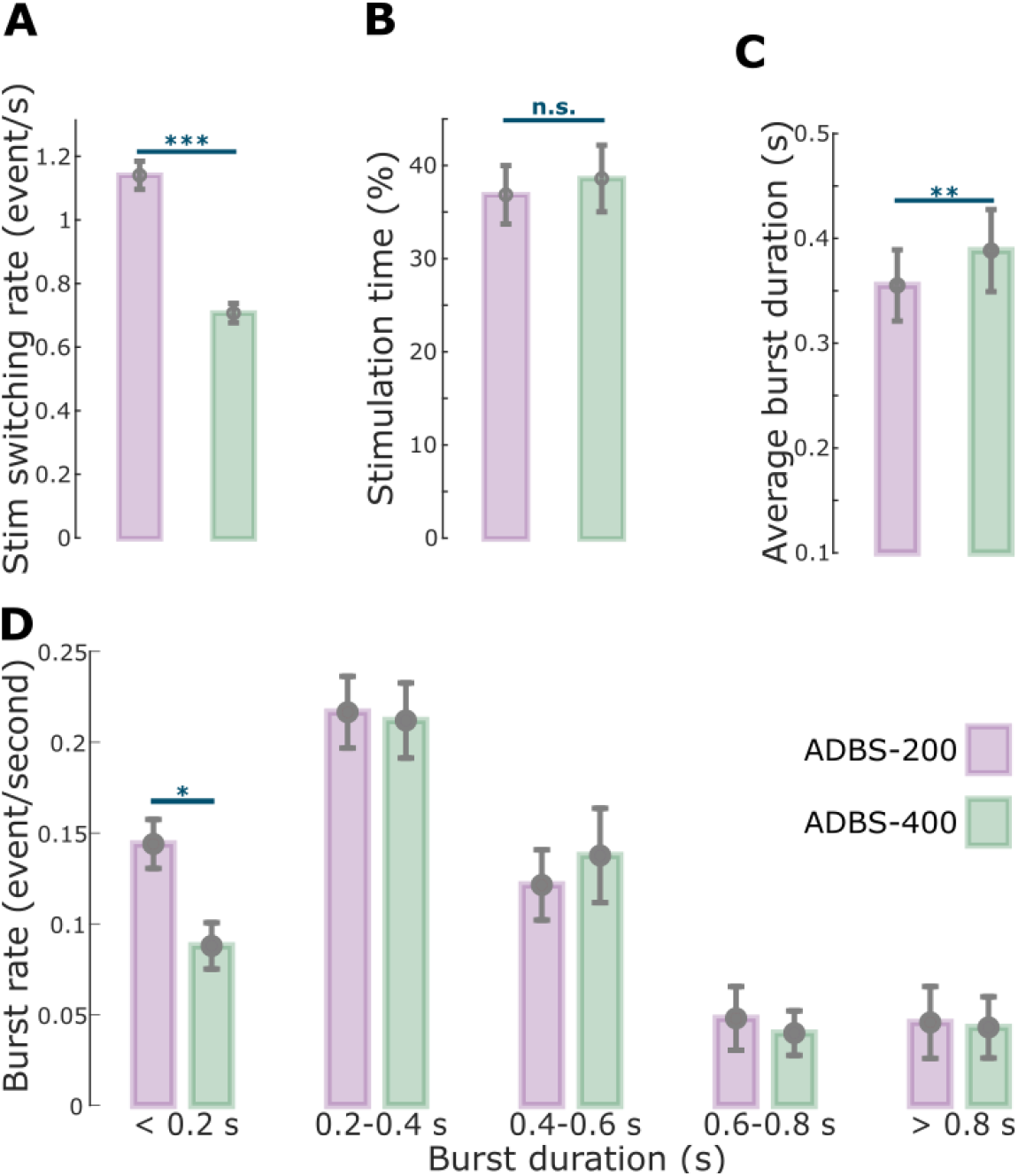
Comparison of the stimulation events and beta bursts between ADBS-200 and ADBS-400 conditions. **(A)-(D)** Averaged stimulation switching rate (A), percentage of time when the stimulation was on (B), averaged duration of beta bursts (C), and averaged rate of beta bursts with different durations (D) in ADBS-200 (purple) and ADBS-400 (green) conditions. The error bar plots show the mean and SEM across all tested hemispheres in different conditions; \**p*<0.05, \*\**p*<0.01; \*\*\**p*<0.001; *p*-values were quantified based on paired t-test on individual hemisphere basis (N=16) and corrected for multiple comparisons using Bonferroni correction.

Despite the clear difference in the stimulator output between ADBS-200 and ADBS-400 as expected from a shorter beta smoothing window, there was no significant difference in motor performance. Specifically, there was no difference in any of the evaluated metrics of the reaching movements including reaction time (*k* = -0.0225 ± 0.0181, *p* = 0.2155, **Fig. 3B**) or mean velocity (Reach: *k* = 0.0065 ± 0.0062, *p* = 0.2977, **Fig. 3C**; Return: *k* = 0.0025 ± 0.0053, *p* = 0.6330, **Fig. 3D**). Similarly, the two ADBS conditions revealed similar performance in the finger-tapping task as evaluated by the root-mean-square acceleration (*k* = 0.0189 ± 0.0310, *p* = 0.5416, **Fig. 3E**) and blinded video ratings (*k* = 0.0047 ± 0.1265, *p* = 0.9703, **Fig. 3F**). There was no difference in resting tremor either (*k* = -0.1683 ± 0.2336, *p* = 0.4714, **Fig. 3G**) between the two ADBS conditions.

**Figure 3.**
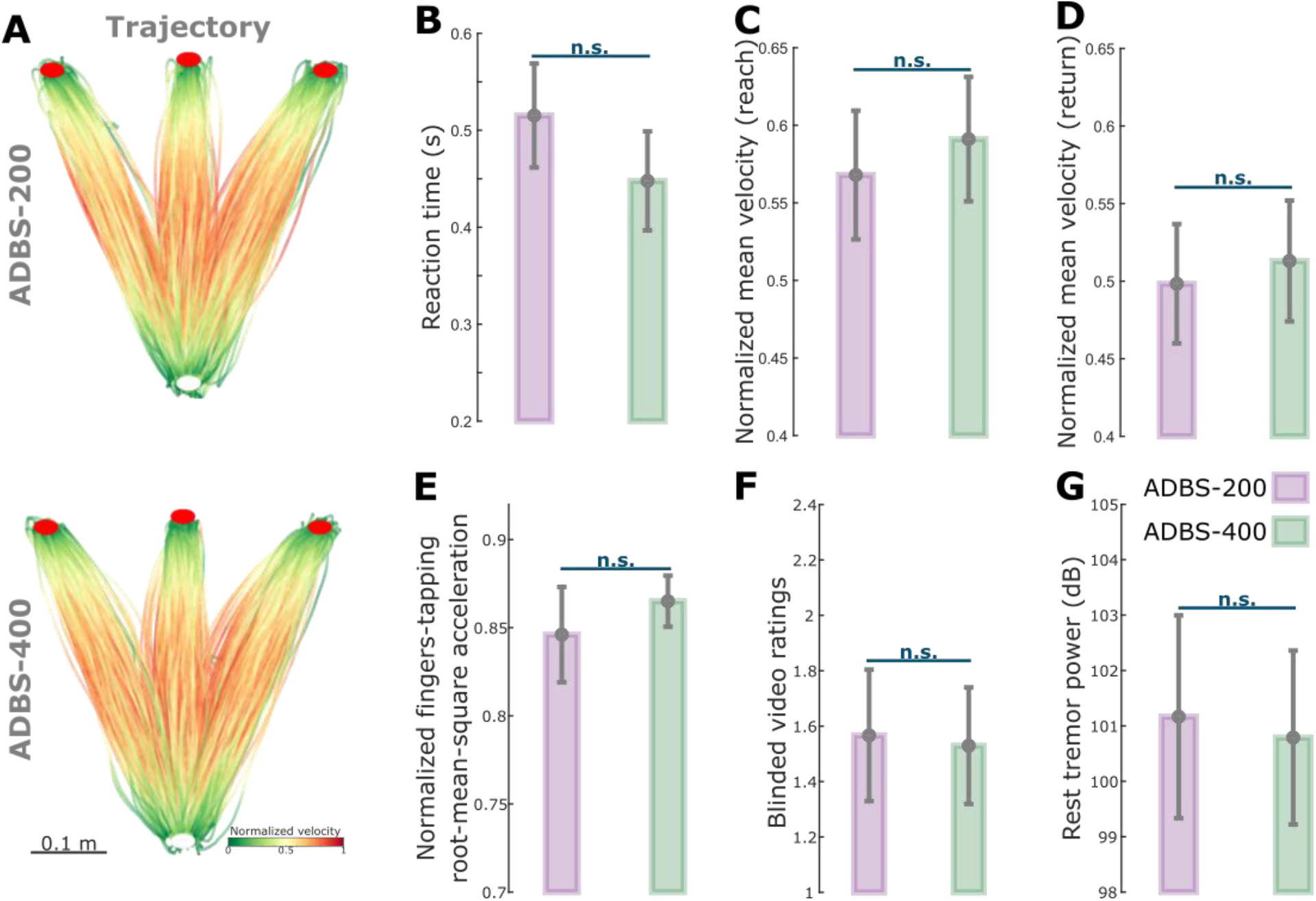
No significant difference in motor performance between ADBS-200 and ADBS-400 conditions. **(A)** Movement trajectories colour coded by the instantaneous velocities of the reaching movement in ADBS-200 (upper) and ADBS-400 (lower) conditions. The velocities were normalized to the individual maximum of each patient. White and red filled circles at the bottom and top indicate the start and target buttons, respectively. **(B)** Reaction time during the reaching movement in different stimulation conditions. **(C)-(D)** Mean velocities during the reaching movement while reach (C) and return (D) periods in different stimulation conditions. **(E)-(F)** Normalized root-mean-square acceleration (E) and blinded video ratings by two experts (F) during finger-tapping movement in different stimulation conditions. **(G)** Average power in tremor frequency band during rest in different stimulation conditions. The error bar plots show the mean and SEM across all tested hemispheres in different conditions. *p*-values were quantified using generalized linear mixed effect modelling on an individual trial (B, C, D, and G) or block (E and F) basis.

Then we compared how these two ADBS conditions modulated the temporal dynamics of beta oscillations. Comparison of the burst characteristics between these two ADBS conditions showed that the average burst duration was shorter during ADBS-200 compared with ADBS-400 (*t*_15_ = -2.9817, *p* = 0.0093, paired *t*-test, **Fig. 2C**). This was mainly due to more bursts with shorter durations (<0.2 s, *t*_15_ = 3.0478, *p* = 0.0081, paired *t*-test, **Fig. 2D**) during ADBS-200, and there was no significant difference in terms of burst rate for bursts with longer durations (>0.2 s) between these two conditions. Please note that here beta bursts were re-quantified offline based on the recorded bipolar LFPs using a 200-ms smoothing window for both ADBS conditions, rather than based on the recorded beta amplitude that had already been smoothed with different time windows for different ADBS conditions. Even though the fast ADBS-200 cut the beta burst even shorter than the ADBS-400, this faster algorithm did not further improve motor performance. These results confirm the findings of previous studies showing that only long beta bursts are pathological.

### 2. ADBS and CDBS equally improved motor performance compared with no DBS, but resting tremor was better suppressed during CDBS

Since we did not see any behavioural difference between ADBS-200 and ADBS-400, we combined these two conditions into one ADBS condition and compared them against CDBS and no DBS for further analysis. Compared with no DBS, both CDBS and ADBS significantly improved motor performance of the cued reaching movements with reduced reaction time (CDBS vs. no DBS: *k* = - 0.0557 ± 0.0217, *p* = 0.0103; ADBS vs. no DBS: *k* = -0.0253 ± 0.0094, *p* = 0.0072, **Fig. 4B**) and increased mean velocity (CDBS vs. no DBS: *k* = 0.0144 ± 0.0058, *p* = 0.0139; ADBS vs. no DBS: *k* = 0.0128 ± 0.0045, *p* = 0.0041, **Fig. 4D**) during backward movements. The effects on the mean velocity during reaching movements were smaller and only significant in ADBS (*k* = 0.0072 ± 0.0028, *p* = 0.0106, **Fig. 4C**) but not in CDBS (*k* = 0.0076 ± 0.0072, *p* = 0.291, **Fig. 4C**) conditions. Both CDBS and ADBS improved the finger-tapping movements with increased root-mean-square acceleration (CDBS vs. no DBS: *k* = 0.0875 ± 0.0372, *p* = 0.0214; ADBS vs. no DBS: *k* = 0.0339 ± 0.0149, *p* = 0.0253, **Fig. 4E**) and reduced blinded bradykinesia ratings (CDBS vs. no DBS: *k* =-0.3088 ± 0.1345, *p* = 0.0249; ADBS vs. no DBS: *k* = -0.1738 ± 0.0593, *p* = 0.0042, **Fig. 4F and Supplementary Video 3**), although some of them were only nominally/marginally significant and did not survive Bonferroni correction for multiple comparisons. When comparing between CDBS and ADBS conditions, no significant behavioural difference was found in any of the evaluated metrics (**Fig. 4B-F**) for the reaching or finger-tapping movements, suggesting that ADBS improved motor performance to a similar extent as CDBS. However, there was more resting tremor during ADBS compared with CDBS (*k* = 0.7605 ± 0.2179, *p* = 0.0005, **Fig. 4G**), even though tremor was significantly reduced in both DBS conditions compared with no DBS (CDBS vs. no DBS: *k* = -2.152 ± 0.3265, *p* = 6.8335e-11; ADBS vs. no DBS: *k* = -0.5726 ± 0.1256, *p* = 5.5933e-06, **Fig. 4G**). The mean duration on stimulation was only 39.39 ± 3.14% of time during ADBS, which was significantly less than CDBS where the stimulation was continuously on (*t*_17_ = 18.1342, *p* = 1.4736e-12, paired *t*-test, **Fig. 4H**).

**Figure 4.**
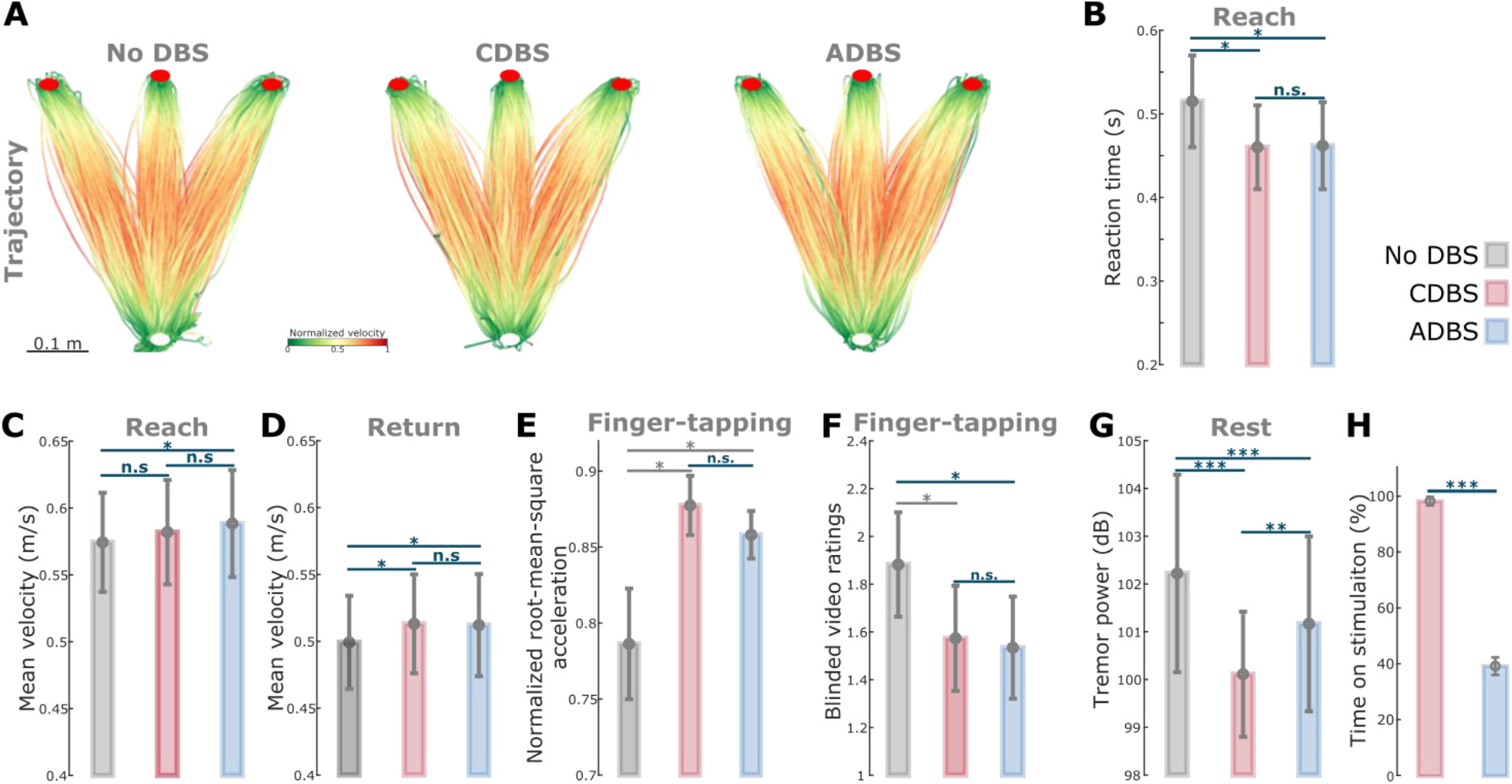
ADBS and CDBS equally improved motor performance compared with no DBS, but resting tremor was better suppressed during CDBS. **(A)** Movement trajectories are colour coded by the normalized instantaneous velocities of the reaching movements with no DBS (left), CDBS (middle), and ADBS (right). White and red filled circles at the bottom and top indicate the start and target buttons, respectively. **(B)** Reaction time during the reaching movement in different stimulation conditions. **(C)-(D)** Mean velocities during the reaching movement while reach (C) and return (D) periods in different stimulation conditions. **(E)-(F)** Normalized root-mean-square acceleration (E) and blinded video ratings (F) during finger-tapping movement in different stimulation conditions. **(G)** Average power in tremor frequency band during rest in different stimulation conditions. **(H)** Time on stimulation in CDBS and ADBS conditions. The error bar plots show the mean and SEM across all tested hemispheres; \**p*<0.05, \*\**p*<0.01; \*\*\**p*<0.001; *p*-values were quantified using generalized linear mixed effect modelling on an individual trial (B, C, D, and G) or block (E and F) basis or using paired t-test on an individual hemisphere basis (H) and corrected for multiple comparisons using Bonferroni correction. Grey * indicates nominally/marginally significant which did not survive Bonferroni correction.

### 3. Stimulation probability during ADBS followed a similar pattern as movement-related beta modulation

To further investigate the modulation of beta and stimulation probability by movement and how these movement-related modulations were changed by different stimulation conditions, we quantified time-frequency spectrograms from bipolar STN LFPs and instantaneous movement velocities, and aligned them to the onset of the reaching movement or the time when the target was reached. During all DBS conditions, a clear event-related desynchronization (ERD) in the beta frequency band (13-30Hz) was observed around onset of the reaching movement (**Fig. 5A-C**), as well as around the time when the target was reached, before the initiation of return movements (**Fig. 5E-G**). In fact, the beta power reached its minimum around both reaching and return movement initiations, then resynchronized to or above baseline level at the end of the movements (**Fig. 5D and H**). During ADBS, the averaged stimulation probability followed a similar pattern as the modulation of beta, but with a constant shift in time that was caused by real-time filtering and smoothing (**Fig. 5D and H**). In general, the stimulation probability was lowest around 0.5 s after the initiation of the reaching movement in this paradigm, with an average stimulation probability of 32.55 ± 4.80% in the 1-s time window.

**Figure 5.**
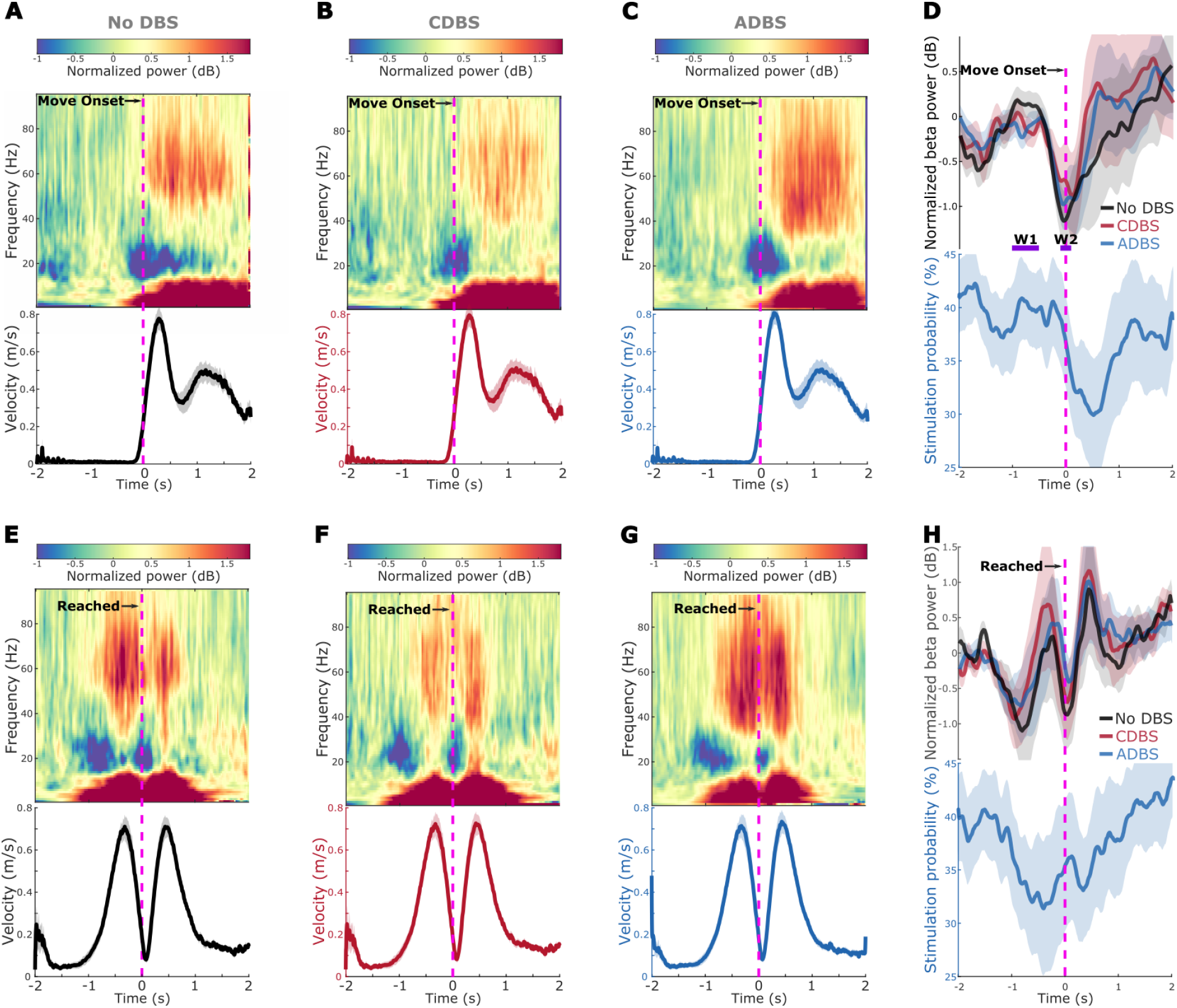
Modulation of beta/gamma power and stimulation probability during reaching movement. **(A-C)** Group averaged time-frequency power-spectra of the targeted STN LFPs aligned to movement onset during reaching movement in no DBS (A), CDBS (B), and ADBS (C) conditions. The power spectra were normalized against a 1-s pre-Go cue resting period in each individual trial. Beta was suppressed around movement initiation and gamma was increased during movement. Lower panel in each subplot indicates the group averaged velocity during the reaching movement. **(D)** Group averaged beta power in different conditions (upper) and stimulation probability during ADBS (lower) aligned to movement onset during reaching movement. Different colors indicate different conditions. Solid line and shade indicate the mean and SEM of the velocity, beta power, or stimulation probability averaged across all hemispheres, respectively. W1 and W2 indicate two time windows where the average beta power was used for predicting reaction time in **Table 1. (E-H)** The same as (A-D) but aligned to the time when the target was reached.

### 4. Reaction time and mean velocity during the reaching movement were predicted by STN beta and gamma power

The spectrograms averaged across trials and time locked to the movement initiation also revealed clear gamma power increase during the execution of reaching movements (**Fig. 5 A-C and E-G**). Previous studies have showed a complementary role of beta desynchronization and gamma increase during movement in invigorating movements (*35*). In particular, gamma power during movements correlated with maximal speed (*36, 37*). Here we further explored the potential associations between beta/gamma oscillations and motor performance, as well as the effect of different DBS protocols. To do so, for each individual trial, we first quantified beta power at different time windows, including average beta power in the 1 to 0.5 s window (W1 in **Fig. 5D**) before movement initiation (*β*_*w*1_) as baseline, average beta power in the 0.2 s window (W2 in **Fig. 5D**) around movement initiation (*β*_*w*2_) where beta was minimal, and beta ERD as the difference between *β*_*w*1_ and *β*_*w*2_. Then, we used each of these beta power windows, together with stimulation condition index (1: no DBS; 2: CDBS or ADBS; Here CDBS and ADBS were combined since there was no behavioural difference between them) as independent variables to predict the reaction time of the reaching movements in separate GLME models. As shown in **Table 1**, the results suggested that although there was a positive estimation effect for *β*_*w*1_ and a negative estimation effect for *β*_*w*2_ in predicting reaction time, neither of the effects was significant. However, there was a significant positive estimation effect on beta ERD (*k* = 0.0301 ± 0.0150, *p* = 0.0453) in predicting reaction time, together with a significant negative estimation effect on stimulation condition (*k* = - 0.0742 ± 0.0363, *p* = 0.0409), suggesting stimulation and smaller beta ERDs independently predicted shorter reaction times. The model also revealed that the interaction between stimulation condition and beta ERD in predicting reaction time was not significant, suggesting that the association between beta ERD and reaction time was not altered by different stimulation conditions. In addition, likelihood ratio test revealed that the GLME model using beta ERD significantly outperformed the model using *β*_*w*1_ (LRStat: 6.748; p < 0.001, chi-squared test) or *β*_*w*2_ (LRStat: 1.8418; p < 0.001, chi-squared test) in predicting reaction time.

**Table 1.**
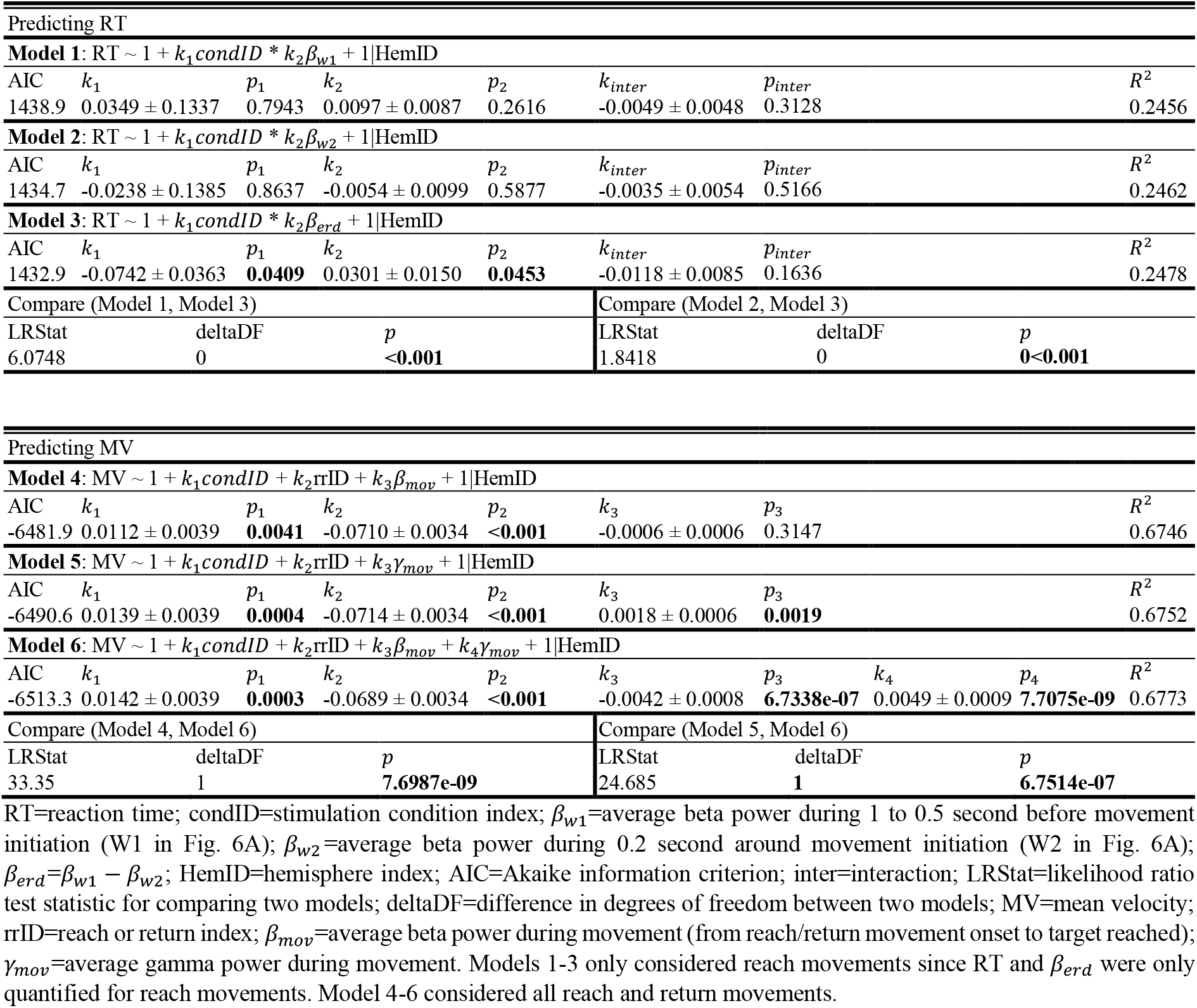
Effects of beta/gamma power in predicting motor performance during reaching movement revealed by generalized linear mixed effect (GLME) modelling.

When using the average beta power during movement, average gamma power during movement, stimulation condition index (1: no DBS; 2: CDBS or ADBS), and reach or return index (1: reach; 2: return) as four independent variables in a GLME model to predict mean velocities, the modelling results revealed significant negative estimation effect on beta power (*k* = -0.0042 ± 0.0008, *p* = 6.7338e-07) and positive estimation effect on gamma power (*k* = 0.0049 ± 0.0009, *p* = 7.7075e-09), suggesting less beta and more gamma during movement together predicted bigger velocities. Apart from this, the modelling also revealed that the mean velocities were bigger during DBS compared with no DBS conditions (*k* = 0.0142 ± 0.0039, *p* = 0.0003), and during reach movements compared with return movements (*k* = -0.0689 ± 0.0034, *p* < 0.001), which were consistent with the results shown in **Fig. 4**. The GLME model combining both beta and gamma performed significantly better than the model only considered beta (LRStat: 33.35; p = 7.6987e-09, chi-squared test) or gamma (LRStat: 24.685; p = 6.7514e-07, chi-squared test) in predicting mean velocities, further confirming that beta and gamma simultaneously associated with the mean velocity during the reaching movement.

### 5. Both STN beta and gamma were overall suppressed by DBS, and the suppression was stronger during CDBS compared with ADBS

Since beta ERD contributed to predicting reaction time and both beta reduction and gamma increase power contributed to predicting movement speed, we investigated the effect of different DBS protocols on these oscillations in the STN to see if we can explain the observed behavioural results that both ADBS and CDBS improved movement speed and reaction time to a similar degree. To do so, we normalized the beta and gamma power in three DBS conditions against the average beta and gamma power during the 1-s pre-Go cue resting period in the no DBS condition, and aligned them to the onset of the reaching movement. As shown in **Fig. 6 A and D**, on top of the movement related modulation, STN beta and gamma power were overall suppressed by DBS, which has been reported in previous studies (*38*), and the suppression was stronger during CDBS compared with ADBS conditions. Specifically, compared with no DBS, the suppression of beta and gamma during CDBS was significant along the whole-time course, while the suppression of beta and gamma during ADBS was only significant at certain time windows. We then compared the averaged beta power in the different time windows used in **Table 1** among different stimulation conditions. The results further confirmed that both ADBS (*β*_*w*1_: *k*=-0.5508 ± 0.0696, *p* = 4.8748e-15, **Fig. 6B**; *β*_*w*2_: *k*=-0.3061 ± 0.0839, *p* = 0.0003, **Fig. 6C**) and CDBS (*β*_*w*1_: *k*=-2.3452 ± 0.1816, *p* = 1.5153e-35, **Fig. 6B**; *β*_*w*2_: *k*=-1.4809 ± 0.1961, *p* = 9.157e-14, **Fig. 6C**) significantly suppressed beta, and the suppression of beta was stronger during CDBS compared with ADBS (*β*_*w*1_: *k*=-1.1832 ± 0.1135, *p* = 1.2901e-24, **Fig. 6B**; *β*_*w*2_: *k*=-0.8398 ± 0.1613, *p* = 2.1741e-07, **Fig. 6C**). In addition, we found beta ERD was also significantly reduced during DBS condition compared with No DBS (CDBS vs. no DBS: *k* = -0.8642 ± 0.2082, *p* = 3.5655e-05; ADBS vs. no DBS: *k* = -0.2439 ± 0.0990, *p* = 0.0138). However, there was no difference between CDBS and ADBS (*k* = 0.3476 ± 0.1932, *p* = 0.0723). Results in the previous section showed that reaction time was more related to beta ERD. The results here may explain why CDBS and ADBS lead to similar changes in reaction time.

**Figure 6.**
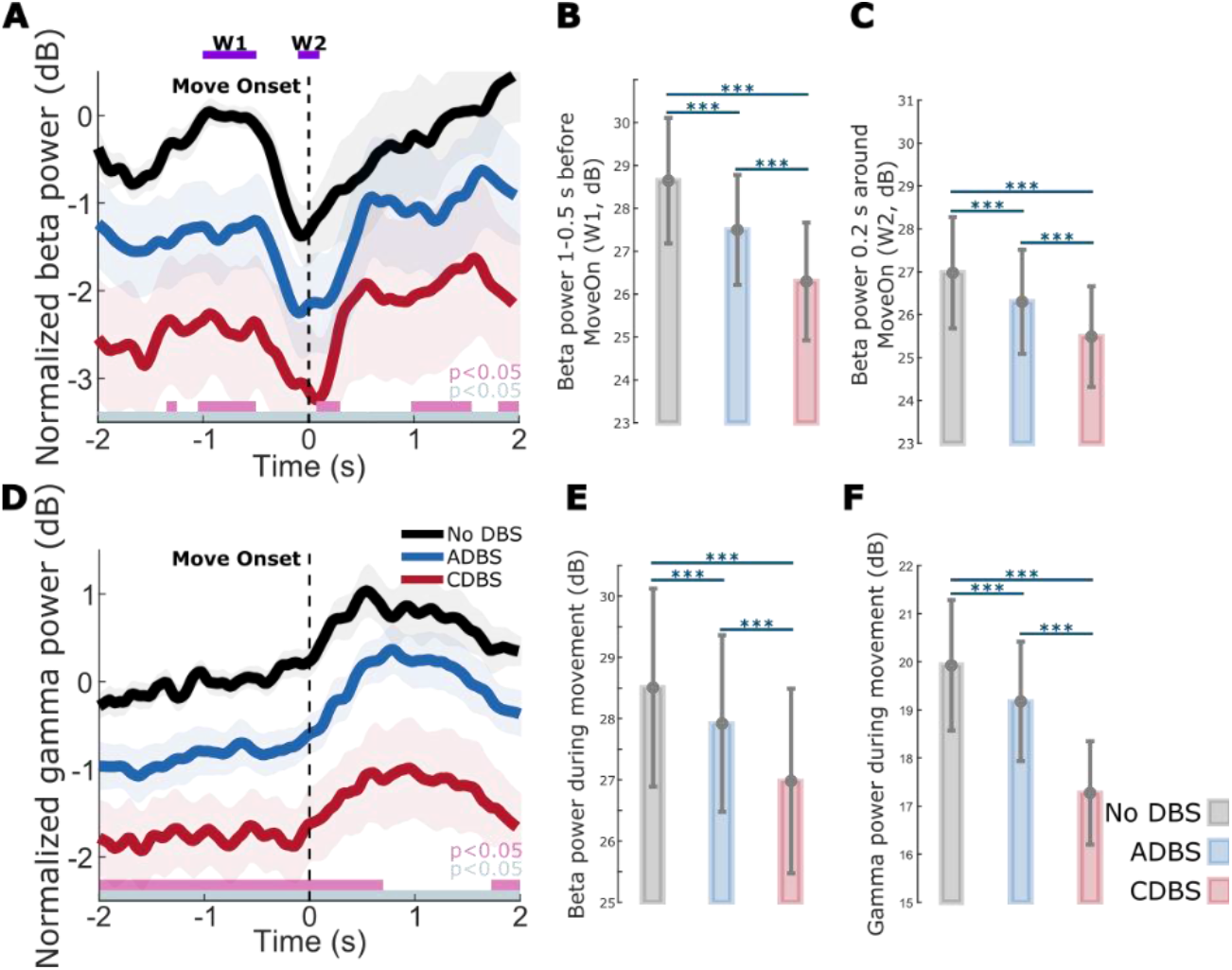
Beta and gamma power were both suppressed during DBS compared with no DBS, and the suppression was stronger during CDBS compared with ADBS. **(A)** Group averaged beta power aligned to movement onset during reaching movement in different conditions. The power was normalized against the average beta power during the 1-s pre-Go cue resting period in no DBS condition. Solid line and shade indicate the mean and SEM of the beta power, respectively. Gray and pink bars on the bottom indicate significant difference between no DBS and CDBS, and between no DBS and ADBS based on a cluster-based permutation procedure, respectively. **(B)-(C)** Averaged beta power without baseline normalization in a baseline time window (W1, 1-0.5 s pre-Onset) (B) and a 0.2-s time window around movement initiation (W2) (C) in different conditions. **(D)** The same as (A) but for gamma power. **(E)-(F)** Averaged beta (E) and gamma (F) power without baseline normalization during movement in different conditions. The error bar plots show the mean and SEM across all tested hemispheres in different conditions; \**p*<0.05, \*\**p*<0.01; \*\*\**p*<0.001; *p*-values were quantified using generalized linear mixed effect modelling on an individual trial basis and corrected for multiple comparisons using Bonferroni correction.

Similarly, beta (ADBS: *k*=-0.2756 ± 0.0749, *p* = 0.0002, **Fig. 6E**; CDBS: *k*=-1.5111 ± 0.1910, *p* = 6.2992e-15, **Fig. 6E**) and gamma (ADBS: *k*=-0.3937 ± 0.0767, *p* = 3.2493e-07, **Fig. 6F**; CDBS: *k*=-2.6497 ± 0.2023, *p* = 1.8303e-36, **Fig. 6F**) power during movement (from reach/return movement onset to target reached) were significantly suppressed by DBS, and the suppression was stronger during CDBS compared with ADBS (Beta: *k*=-0.9337 ± 0.1189, *p* = 7.6942e-15, **Fig. 6E**; Gamma: *k*=-1.8405 ± 0.1074, *p* = 2.3989e-60, **Fig. 6F**). The results here may explain why CDBS and ADBS lead to similar changes in movement speed: even though CDBS suppressed beta more than ADBS, it also suppressed gamma more, whereas both reduction of beta and increase of gamma contributed to invigorating movements.

## Discussion

There were three main findings from this study. First, we showed that shortening the smoothing window to 200ms did make the ADBS more responsive. Further, it shortened the average duration of beta bursts by increasing the number of bursts shorter than 200ms. However, this did not bring any behavioural benefit compared with ADBS with a 400-ms smoothing window for estimating beta, supporting the argument that short beta bursts of less than 400ms can be physiological and there is no benefit from reducing them. Second, we showed that although beta-triggered ADBS reduced the average time on stimulation during reaching movements, it did not compromise motor performance in terms of reaction time and movement speed compared with CDBS. Both ADBS and CDBS improved the performance of reaching and finger-tapping movements to a similar extent compared with no DBS. Third, our results indicated that although ADBS achieved similar effect as CDBS in reducing bradykinesia and improving reaction time and movement speed, it was not as effective as CDBS in suppressing resting tremor.

### Why is there no behavioural difference between ADBS-200 and ADBS-400?

Previous studies showed that STN beta bursts with different durations might have different roles in PD. In particular, the occurrence of longer beta bursts with large amplitude positively correlates with motor impairment (*10-13*). Here, in addition to the commonly used 400-ms smoothing time window (ADBS-400) (*10, 14-16*), we also tested a faster ADBS algorithm in which a 200-ms smoothing time window was used (ADBS-200), to test whether this might further improve the efficacy of ADBS. Our results showed no difference between these two ADBS conditions in any of the evaluated motor performance metrics, including reaction time, movement velocity, resting tremor, root-mean-square acceleration and blinded video ratings of finger-tapping (**Fig. 3**). This was unlikely due to errors in implementation of these two algorithms, as post-hoc analysis confirmed that ADBS-200 was more responsive to the beta oscillations leading to more frequent switching on/off of the stimulator (**Fig. 2A**) despite a similar total stimulation on time (**Fig. 2B**) compared with ADBS-400. We further compared how the two ADBS strategies modulated beta burst characteristics and found that ADBS-200 reduced the average beta burst duration compared with ADBS-400 (**Fig. 2C**), by increasing the number of shorter bursts with durations less than 200ms while keeping a similar number of longer bursts (**Fig. 2D**). These results further support the hypothesis that only long beta bursts (> 400ms) have a pathological effect in PD (*10-13*). Therefore, being more responsive to those short bursts with durations less than 400ms appears unnecessary.

### Why did ADBS provide comparable improvement in motor performance to CDBS?

STN beta-triggered adaptive DBS has been shown to be at least as effective as conventional continuous DBS as evaluated by MDS-UPDRS-III in multiple studies (*8, 14-21*), but it is still unclear whether beta-triggered ADBS is as effective when patients are engaged in a motor task, since STN beta is suppressed during movement initiation and execution (*22-24*). A recent study of three people with PD showed that ADBS might negatively affect the returning part of a reaching movement and delay movement termination (*17*), although motor improvement as measured by MDS-UPDRS-III was comparable to CDBS. In this study, we found that ADBS achieved similar effects as CDBS in improving motor performance in a reaching task in terms of reaction time, movement velocity, and in improving bradykinesia measured by root-mean-square acceleration and blinded video ratings of finger-tapping movements (**Fig. 4**). Therefore ADBS, despite reduced stimulation during ballistic reaching movements (**Fig. 5C-D**), did not appear to compromise movement initiation or execution compared with CDBS.

There are two explanations for this finding. First, even though beta power is reduced during movements when averaged across trials, transient episodes of long beta bursts can still be observed in individual trials (*39*). This explains why in this study, some stimulation (∼30% of the time) was still delivered during movement in the ADBS conditions (**Fig. 5D, H**). We hypothesize that long pathological beta bursts can still occur during movements, which can be curtailed by ADBS, leading to improvement in motor performance. Second, our analysis revealed that during reaching movement, the reaction time was predicted by beta ERD (**Table 1**), which was significantly reduced during DBS compared with no DBS with no difference between CDBS and ADBS. Furthermore, reduced beta power and increased gamma power during movement together predicted faster movement speed (**Table 1**). In terms of movement velocity, previous studies showed that gamma power in the human basal ganglia is positively correlated with movement speed in patients with either PD (*37*) or dystonia (*40*). Here we show that both STN beta and STN gamma power during movement help predict movement speed, with significant negative and positive estimation effects for beta and gamma, respectively. However, both beta and gamma power were more strongly suppressed during CDBS compared with ADBS (**Fig. 6**). This suggests that although beta was better suppressed during CDBS, ADBS preserved gamma better which help invigorate movements, so that the overall movement speeds were similar during CDBS and ADBS conditions.

### Why is ADBS is not as effective as CDBS in suppressing resting tremor?

Previous studies have demonstrated that STN beta oscillations positively correlate with the severity of bradykinesia and rigidity, but not with resting tremor (*4-8, 29, 41-43*). Several existing trials testing the performance of STN beta-triggered ADBS in chronically implanted patients showed re-emergence of tremor during ADBS in some tremor-dominant people with PD, although its effectiveness with bradykinetic phenotypes has been consistently demonstrated (*18, 44*). Indeed, a decrease of beta activity during parkinsonian tremor has been reported in several studies (*45, 46*). In the presence of tremor, neuronal oscillations at tremor frequency (3−7 Hz) tend to increase in the cortical-basal ganglia-thalamic circuit (*47*), whereas beta power (13−30 Hz) and beta band coupling in the motor network are reduced (*45*). Our previous study also showed that in people with PD with pre-existing symptoms of tremor, successful volitional beta suppression through neurofeedback training was associated with an amplification of tremor, which correlated with increased theta band activity in STN LFPs (*32*). These results suggest that the underlying pathophysiology for tremor is different from that for bradykinesia and rigidity in PD. Both CDBS and ADBS significantly improved motor performance and resting tremor compared with no DBS. However, resting tremor was better suppressed during CDBS than ADBS (**Fig. 4**). These results suggest that apart from STN beta, an additional biomarker for resting tremor might be required while developing ADBS strategies for simultaneous control of bradykinesia/rigidity and tremor in PD.

### Remaining challenges for the development of ADBS systems for PD

The results of this study have implications for the further development of ADBS systems for PD. First, we confirmed that tracking the fast beta dynamics using a short smoothing time window does not bring any additional advantage compared to the 400ms windows used in previous trials. More research effort should be invested in addressing the remaining issues of stimulation artefacts and self-triggering related to the fast termination of stimulus trains (*27*). In our study, a 250-ms ramping up/down during each switching on/off plus a 50-ms refractory time after each switching off were utilized to minimize this issue. However, this could be improved at a hardware level (*48*). Alternatively, continuous modulation of the stimulation intensity using proportional control could also remove the self-triggering problem. Second, it might be more beneficial to combine STN beta, gamma, and real-time detection of the patient’s movement status in creating an enhanced adaptive stimulation algorithm. For instance, suppressing beta while minimizing the suppression of gamma during movement might result in improved motor performance, and less stimulation induced adverse events, such as dysarthria, gait disturbances and dyskinesia. Several previous studies have demonstrated the feasibility of detecting movement state based on bioelectrical signals recorded from the cortical-basal ganglia-thalamic circuit in people with PD or essential tremor (*21, 28, 49-51*). However, extracting gamma power in real-time using chronically implanted devices might still be challenging considering stimulation artefacts. In addition, when gamma oscillation is to be used as a feedback signal, movement-related gamma increase, which tends to correlate with movement speed, needs to be differentiated from finely-tuned gamma which might be an indicator of dyskinesia (*52*). Third, additional feedback signal(s) apart from STN beta might be required to develop an ADBS systems for tremor-dominant people with PD.

## Limitations

In this study, we found that beta-triggered ADBS was still as effective as CDBS during reaching and finger-tapping movements. However, it should be noted that all experiments were conducted 3-6 days after the first surgery for DBS electrode implantation, when the postoperative stun effect was appreciable. In addition, the stimulation configurations used in this study, such as ring-mode construction for directional DBS leads, selection of the stimulation contact, amplitude, frequency, pulse width, etc., could be suboptimal and different from what are used in clinical practice. Therefore, the effect of DBS in general could be further improved. However, the same stimulation parameters were used in all tested DBS conditions within each patient, allowing for a fair comparison between the different conditions. Another limitation is that only short-term effects of DBS were considered during two specific motor tasks, i.e., ballistic reaching and finger-tapping movements. It is unclear to what degree the achieved results could be generalised to longer experimental periods, especially when patients are engaging in normal activities of daily living.

## Conclusion

This study evaluated the effectiveness of STN beta-triggered ADBS during a reaching task involving upper-limb movements in thirteen people with PD. We showed that beta-triggered ADBS did not compromise the motor performance of cued reaching movements in terms of reaction time and movement speed compared with CDBS. ADBS and CDBS significantly improved motor performance by similar amounts compared with no DBS. In addition, we demonstrated that using a shorter smoothing window to estimate beta did make ADBS more responsive. It shortened beta burst durations by increasing the number of beta bursts shorter than 200ms, but this did not bring any additional benefit in motor performance. We also showed that both STN beta reduction and gamma power increase during movement helped in predicting movement speed, suggesting that combining beta, gamma and movement status might confer added benefit in ADBS. In addition, beta-triggered ADBS was not as effective as CDBS in suppressing parkinsonian resting tremor, suggesting that additional feedback signals might be required for tremor-dominant patients. These findings have significant implications for the further development of ADBS algorithms to improve the treatment for PD.

## Materials and methods

### Human subjects

Thirteen people with PD (six females) participated to the study after being recruited at two different centres; King’s College Hospital (KCH) and St George’s Hospital (SGH) (clinical details summarised in **Supplementary Table 1**). All underwent bilateral implantation of DBS electrodes targeting the motor area of the STN. The implanted DBS leads were temporarily externalized prior to a second surgery to connect them to a neurostimulator. Lead placements were confirmed by fusion of preoperative MRI and postoperative CT scans, which were further confirmed by reconstructing the electrode trajectories and location of different contacts using the Lead-DBS MATLAB toolbox (version 2.6.0) (*26*), as shown in **Fig. 1D and Supplementary Video 1**. The study was approved by the local ethics committees and all patients provided their informed written consent according to the Declaration of Helsinki. Patients participated in this study had an average age of 62.15 ± 1.58 (mean ± SEM) and a disease duration of 10 ± 1.21 years and showed good response to dopaminergic medication with mean scores of the MDS-UPDRS part III of 37.04 ± 2.95 and 12.42 ± 1.67 for medication OFF and ON, respectively. In this study, all experiments were conducted with the patients off their dopaminergic medication for at least six hours.

### Experimental protocol

The protocol involved two tasks: a cued reaching task performed on a Tablet Drawing Monitor (33 × 57 cm, Artist 22, XP-PEN, Japan) with a stylus pen, and a 20s finger-tapping task. The reaching task was programmed in C# (Visual Studio 2013). As shown in **Fig. 1A**, each trial of the reaching task started with presentation of a white-filled circle at the bottom of the monitor indicating that the patient should bring the pen to the starting position when they were ready (Ready Cue). Once the pen was in the starting position, the circle turned green to indicate that the pen was detected. After a variable delay of 1-2 seconds, a red-filled circle (the Go-cue) appeared on one of the three potential target positions (top-left, top-middle, or top-right of the monitor). Following this Go-cue signal, the patient was instructed to reach the target and come back to the start position as quickly as possible (see **Supplementary Video 2)**. As shown in **Fig. 1B**, the whole experimental session consisted of eight blocks of 15 trials, with an inter-trial interval of 4-5 s (randomised). There were two blocks in each of the four tested stimulation conditions (no DBS, CDBS, ADBS-200, ADBS-400; details in next section). After the reaching movement task, and at the end of each block, the patient was asked to perform finger-tapping movements for 20 s, by tapping their index fingers on their thumbs as wide and fast as possible. An average interval of 67.67 ± 9.20 s (mean ± SEM) was included between two consecutive blocks for washing out the potential stimulation effect from the previous block. The order of the experimental blocks was randomised and counter-balanced across patients. To achieve this, for each patient, the first four blocks included the four stimulation conditions in randomised order, and the four conditions were repeated in reverse order in the second four blocks (Fig 1B).

### Stimulation

Stimulation was applied unilaterally to the hemisphere contralateral to the hand performing the task. The implanted DBS leads were Medtronic non-directional 3389 (2 cases), Medtronic SenSight™ directional with configuration 1-3-3-1 (6 cases), Boston Vercise™ directional with configuration 1-3-3-1 (2 cases), Boston Cartesia X directional with configuration 3-3-3-3-3-1 (1 cases), Boston Cartesia HX directional with configuration 3-3-3-3-1-1-1-1 (1 cases), or Abbott Infinity™ directional with configuration 1-3-3-1 (1 case). For consistency, in cases with directional leads, the segmented contacts were used in ring mode. In cases with Boston Cartesia X/HX directional leads, only the most inferior 4 levels which were supposed to locate in STN were considered for analysis. A highly configurable custom-built neurostimulator certified by the University of Oxford, UK (an improved version based on what was used in *14, 15*) was used to deliver constant current stimulation in monopolar mode. One of the two contacts in the middle was used as the stimulation contact, and an electrode patch attached to the back of the patient was used for reference (**Fig. 1C**). The stimulation had a fixed frequency of 130 Hz, a biphasic pulse width of 60 microseconds, and an interphase gap of 20 microseconds. Four different stimulation conditions were considered in this study, including no DBS, continuous DBS (CDBS), adaptive DBS with the stimulator controlled by the beta amplitude estimated in real-time using a 200-ms smoothing window (ADBS-200), and adaptive DBS with a 400-ms smoothing window (ADBS-400). Before smoothing, the bipolar LFPs were filtered at the selected beta frequency band and rectified (*10, 14, 15*). The implementation of ADBS was the same as in previous studies, apart from using an advanced stimulator and adding a new condition with shorter smoothing windows (ADBS-200) to capture faster beta dynamics. To mitigate transient effects resulting in a re-entrant stimulation loop during ADBS (*27*), ramping was applied at the start and end of each stimulation switching event, which forced the stimulation amplitude to linearly increase to the desired value or decrease to zero within 250ms. In addition, a refractory time window of 50ms was set after stimulation was switched off.

### Selecting stimulation contact and amplitude, and the beta frequency band for feedback

We followed a similar procedure used in previous studies (*14, 28*) to select the stimulation contact and amplitude. Specifically, we delivered continuous DBS to one of the middle two contacts initially at 0.5 mA. We then progressively increased the amplitude by 0.5 mA increments, until clinical benefit was seen without side effects such as paraesthesia, or until 3.5 mA was reached as the maximum amplitude. If no apparent clinical effect was observed, we repeated this procedure for the other middle contact level. Once the stimulation contact and amplitude were selected, a period of 2 minutes of rest recordings were performed. LFPs were recorded from two contacts neighbouring the selected stimulating contact in the differential bipolar mode. To select the individualized beta frequency band for feedback, the recorded LFPs were first notch-filtered at 50 Hz and band-pass filtered between 1 and 95 Hz using a second order zero-phase digital filter. The periodogram power spectral density (PSD) was then estimated. The feedback beta frequency band was selected as ± 3 Hz around the largest beta peak (13-30 Hz). In the ADBS conditions, the threshold for triggering the stimulation was set manually for each hemisphere separately so that the DBS would be switched on for about 50 percent of the time when the patient was at rest (**Fig. 1C**), as in the previous ADBS studies (*8, 10, 14, 15, 18, 29*). For patients who performed the tasks with both hands, the stimulation contact and amplitude, as well as the beta frequency band and triggering threshold were selected separately for each hemisphere. These stimulation parameters (summarized in **Table 2**) were kept constant for different stimulation conditions for each hemisphere.

**Table 2.** Details of the stimulation used during the recording of this study.

| Case | DBS lead | Stim contact (L/R) | Stim Amp (L/R mA) | Bipolar feedback channel (L/R) | Online filter range (L/R Hz) |
| --- | --- | --- | --- | --- | --- |
| 1 | Medt1 | L3 | 3 | L24 | 19-25 |
| 2 | Medt1 | L3 / R2 | 3.5 / 1.5 | L24 / R13 | 14-20 / 15-21 |
| 3 | Bost1 | L2 / R3 | 3 / 2 | L13 / R24 | 15-21 / 14-20 |
| 4 | Bost2 | L3 | 1 | L24 | 16-22 |
| 5 | Abbo | R3 | 1.5 | R24 | 17-23 |
| 6 | Medt2 | R2 | 1.5 | R13 | 19-25 |
| 7 | Bost3 | L2 / R2 | 2.5 / 2.5 | L13 / R13 | 16-22 / 22-28 |
| 8 | Medt2 | R2 | 3 | R13 | 15-21 |
| 9 | Medt2 | L3 | 1.5 | L24 | 14-20 |
| 10 | Medt2 | L2 / R2 | 1 / 3 | L13 / R13 | 22-28 / 22-28 |
| 11 | Medt2 | L3 / R2 | 3.5 / 3.5 | L24 / R13 | 18-24 / 17-23 |
| 12 | Medt2 | L2 / R2 | 3 / 3 | L13 / R13 | 12-18 / 21-27 |
| 13 | Bost1 | L2 / R2 | 2 / 2 | L13 | 18-24 / 20-26 |
| Mean |  |  | 2.38 |  | 17.3-23.3 |
| SEM |  |  | 0.18 |  | 0.66 |

DBS=deep brain stimulation; Stim=stimulation; L=left; R=right; Amp=amplitude; Medt1=Quadripolar non-directional Macroelectrode, Model 3389, Medtronic; Bost1=Vercise™ directional lead with 1-3-3-1 configuration, Boston Scientific; Bost2=Cartesia™ X leads with 3-3-3-3-1 configuration, Boston Scientific; Bost3=Cartesia™ HX leads with 3-3-3-3-1-1-1 configuration, Boston Scientific; Abbo=St. Jude Medical Infinity 0.5mm spaced directional DBS leads with 1-3-3-1 configuration, Abbott; Medt2=SenSight™ 0.5mm spaced directional lead with 1-3-3-1 configuration, Medtronic; SEM=standard error of the mean.

### Data recording

All recordings were carried out 3-6 days after the first surgery for DBS electrode implantation. A TMSi Porti or Saga amplifier (TMS International, the Netherlands) was used to record bipolar LFPs from the two contacts adjacent to the stimulating contact (**Fig. 1C**) at a sampling rate of 2048 Hz (cases 1-2, 4-8, Porti amplifier) or 4096 Hz (cases 3, 9-13, Saga amplifier). The acceleration of the patient moving their hand was measured using a triaxial accelerometer taped to the back of the index finger and simultaneously recorded with the same amplifier at the same sampling frequency as the LFP signals. The precise timing of all cue signals of the reaching task (Start, Go, Reached and Back, **Fig. 1A**) and the finger tapping task (Start/Stop) were captured using a photodiode taped to the monitor, and recorded with the same amplifier. Furthermore, the instantaneous stimulation amplitude applied during the real-time experiment was also simultaneously recorded by a custom-developed C program. The ground electrode was placed on the resting forearm of the patient. The X and Y coordinates of the stylus on the monitor and the corresponding timestamps were recorded automatically at an irregular sampling rate of 84.3062 ± 3.3060 Hz (mean ± SEM) by a custom-developed C# program (irregularity of sampling was due to the imprecision of the timer in C#). In addition, videos of the finger-tapping movements were recorded using a smartphone (iPhone 6s, Apple Inc., US) for further blinded assessment. Among the 13 patients, seven (cases 2-3, 7, 10-13) performed the task with both hands separately, resulted in 20 hemispheres in total. However, the left hemisphere for case 2 was excluded due to strong stimulation artefact contaminating the estimated beta in all stimulation conditions, probably due to the high amplitude of stimulation (3.5 mA) and/or high electrode impedance. Case 5 was excluded due to obvious stimulation induced dyskinesia even at low stimulation amplitude (1.5 mA). The data from the remaining 12 patients (18 hemispheres) were analysed. Due to limited time for conducting the experiment, case 10 did not perform the task in the ADBS-200 condition.

### Kinematic data analysis

#### Reaching movements

The trajectories of the reaching movements were re-constructed for each trial, based on the recorded XY coordinates and timestamps, as shown in **Fig. 3A and Fig. 4A**. The mean velocities of the reach and return movements were calculated separately for each trial by dividing the accumulated distances against the durations of the movements. Instantaneous velocity was quantified using two adjacent coordinates and their timestamps. In addition, the reaction time was defined as the time from the Go-cue to the first timestamp when the pen moved out of the target button.

#### Resting tremor

To investigate the impact of beta-triggered ADBS on resting tremor, we quantified tremor power using the accelerometer measurements recorded from the tested hand 5 s before the Go-cue, when the patient was at rest. More specifically, the recorded three-axes accelerometer signals were first band-pass filtered between 1 and 95 Hz and band-stop filtered between 48 and 52 Hz using 3-order zero-phase Butterworth filters, then decomposed into time-frequency domain using continuous Morlet wavelet transformation with 6 cycles, and a linear frequency scale ranging from 1 to 10 Hz at 0.5 Hz resolution. The average power in tremor frequency band (i.e., 3-7 Hz) across three axes were quantified (10log10 transferred to dB) as the resting tremor severity.

#### Finger-tapping

For each finger-tapping movement, we quantified the root-mean-square acceleration based on the recorded three-axes accelerometer signals as an overall evaluation of the tapping performance (*30, 31*). The root-mean-square acceleration was quantified for each individual block and then normalized against the maximum across all blocks within the same hand. In addition, two experienced movement disorder specialists (F.B. and A.M. in the author list) reviewed the recorded videos of the finger-tapping movements and separately rated the movement according to adjusted MDS-UPDRS-III (finger tapping instruction). The assessors were blinded to the stimulation condition of each video. The mean rating from the two assessors was then calculated. Due to obvious fatigue effect after long-lasting finger-tapping, only the first 10 s accelerometer measurements and video recordings were considered during the assessment.

### Stimulation and LFP data analysis

During ADBS-200 and ADBS-400, the average percentage of time when the stimulation was on and stimulation switching rate (number of stimulation events per second) were quantified based on the recorded stimulation amplitude. Please note that, the stimulation ON time did include the ramping up/down time.

The effects of the two different ADBS algorithms on the dynamics of the beta oscillations were also analysed. The bipolar LFPs recorded from the feedback channel for each task were processed off-line in the same way as used for real-time beta estimation, with the only difference that a 200-ms smoothing window was used for all conditions, so that we could compare dynamics of beta oscillations across stimulation conditions. Then the 75th percentile of the beta amplitude with the patient at rest and stimulation off was used to define beta bursts. Next, average burst duration and burst rate (events per second) were quantified as described before (*10, 32*). To investigate the movement related modulation in the STN, LFPs were first epoched starting 5 seconds before the Go-cue to 2 seconds after the pen returned to the start button. Then the signals were band-pass filtered between 1 and 95 Hz, band-stop filtered between 48 and 52 Hz using 3-order zero-phase Butterworth filters, and decomposed into time-frequency domain using continuous Morlet wavelet transformation with a linear frequency scale ranging from 1 to 95 Hz at 1 Hz resolution, and a linearly spaced number (4−8) of cycles across all calculated frequencies. The calculated power of each time point at each frequency was decibel (dB) baseline normalized against the average power in the 1 second window before the Go-cue. The beta and gamma power were also quantified as the average power in the frequency bands of 13-30 Hz and 35-90 Hz, respectively.

### Statistical analysis

Statistical analyses were conducted using custom-written scripts in MATLAB R2021-b (The MathWorks Inc, Nantucket, Massachusetts).

For those metrics quantified on per condition basis (including stimulation switching rate, average time when the stimulation was on, average burst duration, and burst rate), paired t-tests were used to evaluate the effect of the stimulation condition. The normal distribution assumption was tested using Anderson-Darling test. Multiple comparisons applied to different measurements were corrected using Bonferroni correction.

For those metrics quantified on individual trial/block basis (including reaction time, mean velocity, rest tremor power, root-mean-square acceleration, and blinded video rating), generalized linear mixed effect modelling (GLME) (*33*) was used to investigate the effect of different stimulation conditions. Due to the naturally skewed characteristic of reaction time, normal distribution with log link function was used in the models using reaction time as the dependent variable. Otherwise, normal distribution with identity link function was used. We also used GLME to further investigate the effects of STN beta/gamma power on performance of the reaching movement measured by reaction time and mean velocity on a trial-by-trial basis. In each model, the slope(s) between the predictor(s) and the dependent variable were set to be fixed across all hemispheres while a random intercept was set to vary by hemisphere. For each GLME model, the parameters were estimated based on maximum likelihood using Laplace approximation, the Akaike information criterion (AIC), estimate value with standard error of the coefficient (*k* ± *SE*), *p*-value (*p*), and proportion of variability in the response explained by the fitted model (*R*^2^) were reported. A chi-squared reference distribution based likelihood ratio test was conducted for the comparison of two fitted GLME models, and the likelihood ratio test statistic (LRStat), difference in degrees of freedom between two models (deltaDF), and *p*-value for the likelihood ratio test were reported for each pair of models comparison. The modelling is further detailed below together with the results.

To compare the group averaged beta/gamma power at different time points relative to movement, a nonparametric cluster-based permutation procedure (repeated 1000 times) was applied and multiple comparisons were controlled (*34*).

## Data Availability

The data and codes will be shared on the data sharing platform of the MRC Brain Network Dynamics
Unit.

https://data.mrc.ox.ac.uk/mrcbndu/data-sets/search

## Data availability

The data and codes will be shared on the data sharing platform of the MRC Brain Network Dynamics Unit: https://data.mrc.ox.ac.uk/mrcbndu/data-sets/search.

## Acknowledgement

This work was supported by the Medical Research Council (MC_UU_00003/2) and the BRAIN Non-Clinical Post-Doctoral Fellowship (HMR04170). We thank the participating patients for making this study possible. We thank Andrew O’Keeffe, Natasha Hulse, and Rahul Shah for help with the recordings. We thank Hayriye Cagnan and the rest of the Tan group for providing useful discussions on data analysis.

---

**Supplementary Table 1.**
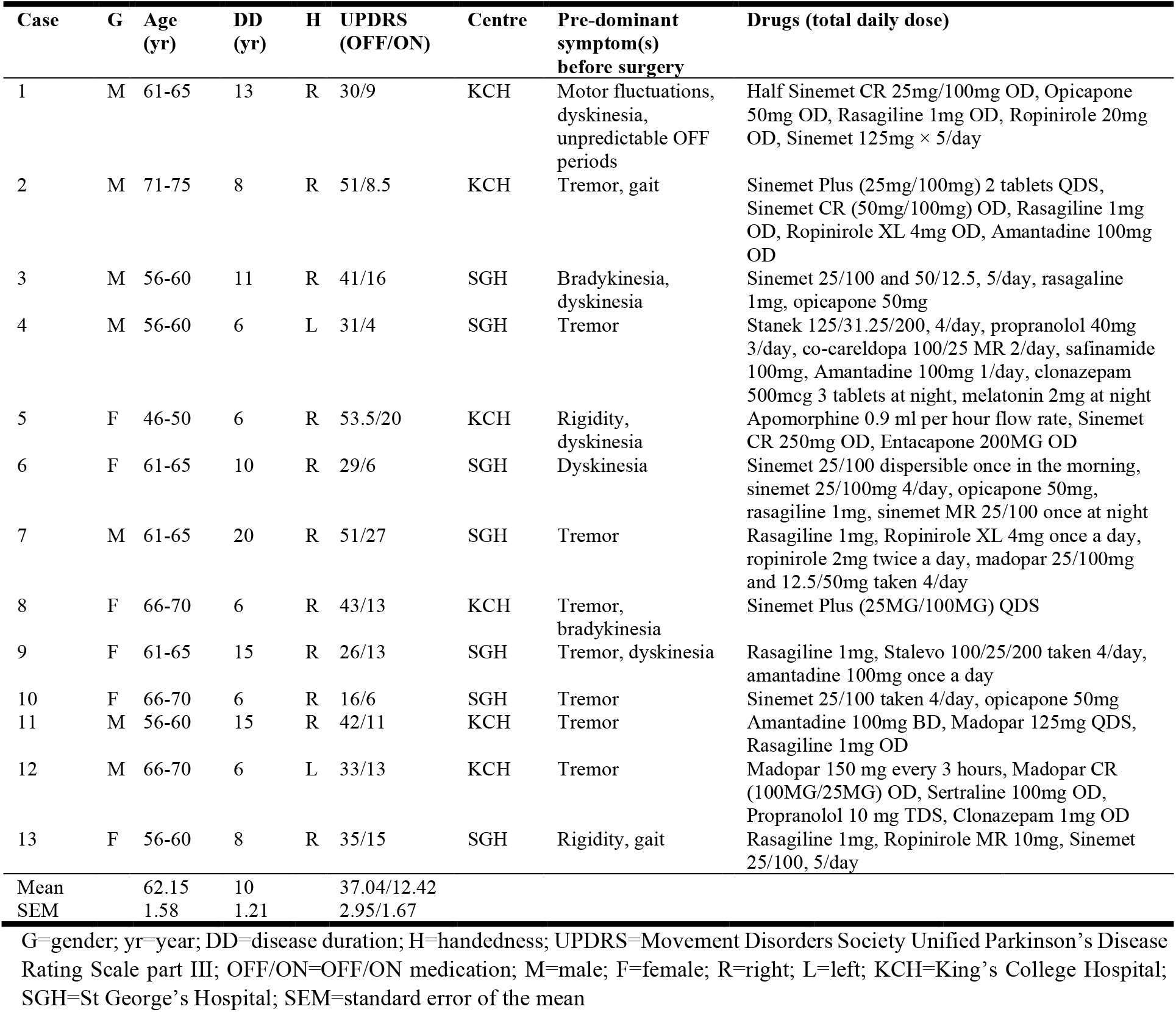
Clinical details of all recorded participants.

| Case | G | Age (yr) | DD (yr) | H | UPDRS (OFF/ON) | Centre | Pre-dominant symptom(s) before surgery | Drugs (total daily dose) |
| --- | --- | --- | --- | --- | --- | --- | --- | --- |
| 1 | M | 61-65 | 13 | R | 30/9 | KCH | Motor fluctuations, dyskinesia, unpredictable OFF periods | Half Sinemet CR 25mg/100mg OD, Opicapone 50mg OD, Rasagiline 1mg OD, Ropinirole 20mg OD, Sinemet 125mg × 5/day |
| 2 | M | 71-75 | 8 | R | 51/8.5 | KCH | Tremor, gait | Sinemet Plus (25mg/100mg) 2 tablets QDS, Sinemet CR (50mg/100mg) OD, Rasagiline 1mg OD, Ropinirole XL 4mg OD, Amantadine 100mg OD |
| 3 | M | 56-60 | 11 | R | 41/16 | SGH | Bradykinesia, dyskinesia | Sinemet 25/100 and 50/12.5, 5/day, rasagiline 1mg, opicapone 50mg |
| 4 | M | 56-60 | 6 | L | 31/4 | SGH | Tremor | Stanek 125/31.25/200, 4/day, propranolol 40mg 3/day, co-careldopa 100/25 MR 2/day, safinamide 100mg, Amantadine 100mg 1/day, clonazepam 500mcg 3 tablets at night, melatonin 2mg at night |
| 5 | F | 46-50 | 6 | R | 53.5/20 | KCH | Rigidity, dyskinesia | Apomorphine 0.9 ml per hour flow rate, Sinemet CR 250mg OD, Entacapone 200MG OD |
| 6 | F | 61-65 | 10 | R | 29/6 | SGH | Dyskinesia | Sinemet 25/100 dispersible once in the morning, sinemet 25/100mg 4/day, opicapone 50mg, rasagiline 1mg, sinemet MR 25/100 once at night |
| 7 | M | 61-65 | 20 | R | 51/27 | SGH | Tremor | Rasagiline 1mg, Ropinirole XL 4mg once a day, ropinirole 2mg twice a day, madopar 25/100mg and 12.5/50mg taken 4/day |
| 8 | F | 66-70 | 6 | R | 43/13 | KCH | Tremor, bradykinesia | Sinemet Plus (25MG/100MG) QDS |
| 9 | F | 61-65 | 15 | R | 26/13 | SGH | Tremor, dyskinesia | Rasagiline 1mg, Stalevo 100/25/200 taken 4/day, amantadine 100mg once a day |
| 10 | F | 66-70 | 6 | R | 16/6 | SGH | Tremor | Sinemet 25/100 taken 4/day, opicapone 50mg |
| 11 | M | 56-60 | 15 | R | 42/11 | KCH | Tremor | Amantadine 100mg BD, Madopar 125mg QDS, Rasagiline 1mg OD |
| 12 | M | 66-70 | 6 | L | 33/13 | KCH | Tremor | Madopar 150 mg every 3 hours, Madopar CR (100MG/25MG) OD, Sertraline 100mg OD, Propranolol 10 mg TDS, Clonazepam 1mg OD |
| 13 | F | 56-60 | 8 | R | 35/15 | SGH | Rigidity, gait | Rasagiline 1mg, Ropinirole MR 10mg, Sinemet 25/100, 5/day |
| Mean |  | 62.15 | 10 |  | 37.04/12.42 |  |  |  |
| SEM |  | 1.58 | 1.21 |  | 2.95/1.67 |  |  |  |

G=gender; yr=year; DD=disease duration; H=handedness; UPDRS=Movement Disorders Society Unified Parkinson's Disease Rating Scale part III; OFF/ON=OFF/ON medication; M=male; F=female; R=right; L=left; KCH=King's College Hospital; SGH=St George's Hospital; SEM=standard error of the mean

**Supplementary Video 1: Lead-DBS Scene**

**Supplementary Video 2: Reaching task (P4 right hand)**

**Supplementary Video 3: Finger-tapping task (P11 right hand)**

